# Associations of comorbidities and medications with COVID-19 outcome: A retrospective analysis of real-world evidence data

**DOI:** 10.1101/2020.08.20.20174169

**Authors:** Basel Abu-Jamous, Arseni Anisimovich, Janie Baxter, Lucy Mackillop, Marcela P. Vizcaychipi, Alex McCarthy, Rabia T. Khan

**Author notes:** Joint first author (Basel Abu-Jamous, Arseni Anisimovich). Corresponding author Correspondence address Rabia Tahir Khan Sensyne Health plc, Schrodinger Building, Heatley Road, Oxford Science Park, Oxford, OX4 4GE.

## Abstract

**Background:** Hundreds of thousands of deaths have already been recorded for patients with the severe acute respiratory syndrome coronavirus (SARS-CoV-2; aka COVID-19). Understanding whether there is a relationship between comorbidities and COVID-19 positivity will not only impact clinical decisions, it will also allow an understanding of how better to define the long-term complications in the groups at risk. In turn informing national policy on who may benefit from more stringent social distancing and shielding strategies. Furthermore, understanding the associations between medications and certain outcomes may also further our understanding of indicators of vulnerability in people with COVID-19 and co-morbidities.

**Methods:** Electronic healthcare records (EHR) from two London hospitals were analysed between 1^st^ January and 27^th^ May 2020. 5294 patients presented to the hospitals in whom COVID status was formally assessed; 1253 were positive for COVID-19 and 4041 were negative. This dataset was analysed to identify associations between comorbidities and medications, separately and two outcomes: (1) presentation with a COVID-19 positive diagnosis, and (2) inpatient death following COVID-19 positive diagnosis. Medications were analysed in different time windows of prescription to differentiate between short-term and long-term medications. All analyses were done with controls (without co-morbidity) matched for age, sex, and number of admissions, and a robustness approach was conducted to only accept results that consistently appear when the analysis is repeated with different proportions of the data.

**Results:** We observed higher COVID-19 positive presentation for patients with hypertension (1.7 [1.3-2.1]) and diabetes (1.6 [1.2-2.1]). We observed higher inpatient COVID-19 mortality for patients with hypertension (odds ratio 2.7 [95% CI 1.9-3.9]), diabetes (2.2 [1.4-3.5]), congestive heart failure (3.1 [1.5-6.4]), and renal disease (2.6 [1.4-5.1]). We also observed an association with reduced COVID-19 mortality for diabetic patients for whom anticoagulants (0.11 [0.03-0.50]), lipid-regulating drugs (0.15 [0.04-0.58]), penicillins (0.20 [0.06-0.63]), or biguanides (0.19 [0.05-0.70]) were administered within 21 days after their positive COVID-19 test with no evidence that they were on them before, and for hypertensive patients for whom anticoagulants (0.08 [0.02-0.35]), antiplatelet drugs (0.10 [0.02-0.59]), lipid-regulating drugs (0.15 [0.05-0.46]), penicillins (0.14 [0.05-0.45]), or angiotensin-converting enzyme inhibitors (ARBs) (0.06 [0.01-0.53]) were administered within 21 days post-COVID-19-positive testing with no evidence that they were on them before. Moreover, long-term antidiabetic drugs were associated with reduced COVID-19 mortality in diabetic patients (0.26 [0.10-0.67]).

**Conclusions:** We provided real-world evidence for observed associations between COVID-19 outcomes and a number of comorbidities and medications. These results require further investigation and replication in other data sets.

## Introduction

As of the 16^th^ of August 2020, the COVID-19 pandemic has resulted in more than 770,000 deaths worldwide over the course of a few months. Although most of the confirmed cases of COVID-19 infection show mild or no symptoms, a global concern exists in ensuring that healthcare systems can cope with those that require hospitalisation, especially with the coming winter pressure and while preparing for a potential second wave. Correct identification of individuals with higher risk of presentation or of death due to COVID-19 will not only help hospitals better identify those in need of hospitalisation but will also assist the community in identifying the vulnerable who require more careful shielding.

Hypertension and diabetes have been shown to be associated with poorer outcome in COVID-19. (Perico, et al., 2020). Furthermore, it has been postulated that certain medications may modulate the immune response to COVID-19 infection, positively or negatively.

We have set out to investigate the association of comorbidities and medications with either presentation with COVID-19 positive diagnosis or death due to COVID-19 in a real-world-evidence (RWE) dataset.

## Methods

### The dataset

Electronic health record (EHR) data were obtained from the Chelsea & Westminster Hospital NHS Foundation Trust for 5294 patients presented to the Trust’s hospitals between 1 ^st^ January and 27^th^ May 2020. The patient population consisted of 1253 COVID-19 positive and 4041 COVID-19 negative patients as diagnosed using viral PCR from swap tests.

For these 5294 patients the dataset included patients’ history of primary and secondary diagnoses recorded in the hospital (from 2004 to 2020) medications administered or prescribed in the hospital (from 2010 to 2020), and death status.

### Association analysis of comorbidities with COVID-19 presentation and mortality

We examined the association of comorbidities with COVID-19 positive presentation as well as with inpatient mortality of COVID-19 patients. This is realised by using Fisher’s exact test to compare the outcomes of patients with a given comorbidity (the active arm) and patients without that comorbidity (the control arm). To account for possible confounders, the two arms were matched on age, sex, and the number of admissions of patients in their available history (Figure 1 (b)). To reduce false discoveries, multiple hypothesis testing was conducted using the Benjamini Hochberg (BH) method. Furthermore, a robustness analysis was carried out by running the same test experiment once with 100% of the available data and 20 more times with randomly selected proportions (60%, 70%, 80%, or 90%) of the data. An association is considered significant if its adjusted p-value was smaller than 0.05 in more than 70% of these individual experiments (Figure 1 (a)).

**Figure 1.**
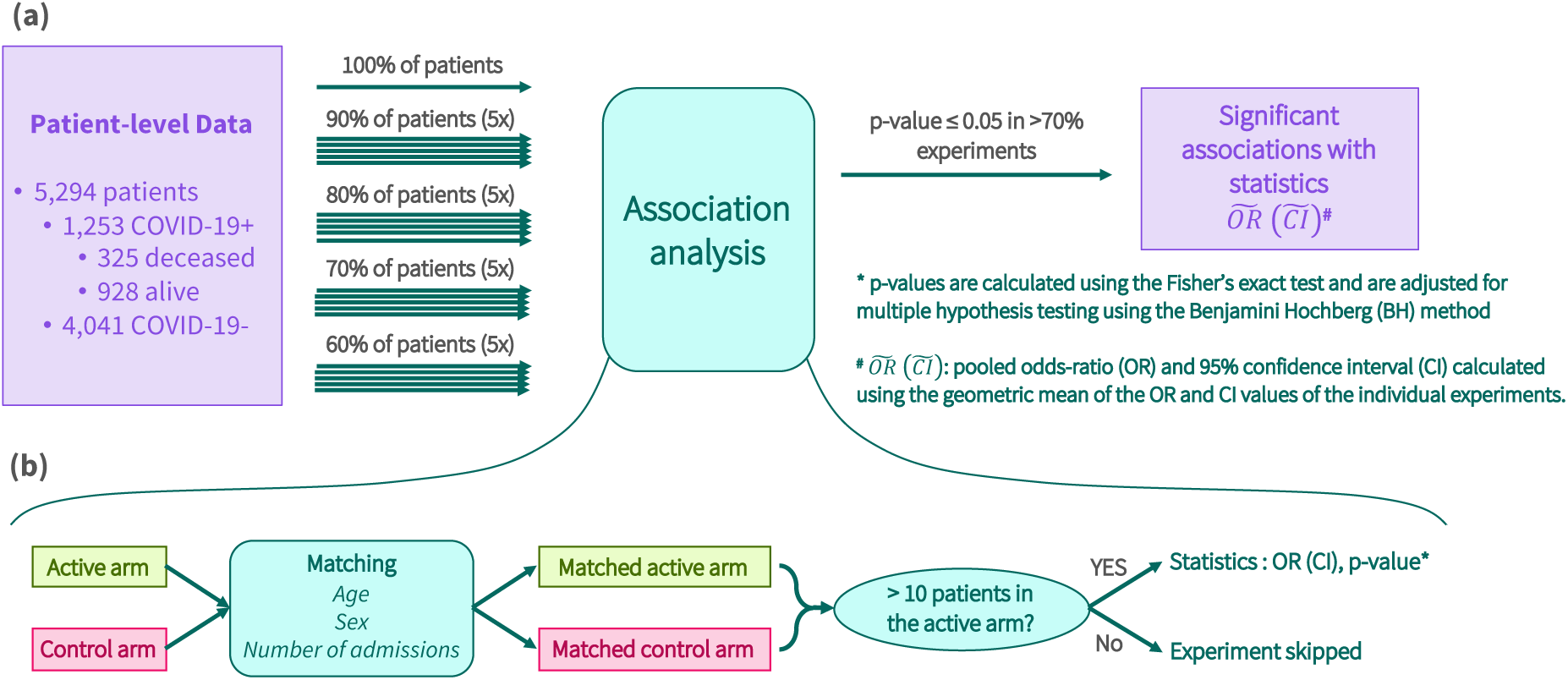
Statistical analysis pipeline. (a) For each tested comorbidity or medication, a total of 21 tests of association are conducted with different proportions of the dataset. The tested association is considered significant if p <= 0.05 in more than 70% of the tests. (b) Each line of analysis is conducted on active and control arms that are matched on age, sex, and number of admissions. Odds-ratios (OR) and 95% confidence intervals (CI) are calculated only if the active arm included more than 10 patients; otherwise, the test yields no result.

Patients are included in the active arm if a record of the given disease appears at least once in their available history. Disease diagnoses are recorded in our data using the International Classification of Diseases, tenth revision (ICD-10) codes. ICD-10 codes were translated into 19 disease groups following the approach proposed by (Quan, et al., 2005) with some amendments ensuring that hypertension and asthma were individually assessed in the analysis (Table 1).

**Table 1.** ICD-10 codes used for comorbidity grouping from (Quan, et al., 2005)^*^

| Disease group | ICD-10 codes |
| --- | --- |
| Myocardial infarction | I21, I22, I252 |
| Congestive heart failure | I43, I50, I099, I110, I130, I132, I255, I420, I425, I426, I427, I428, I429, P290 |
| Peripheral vascular disease | I70, I71, I731, I738, I739, I771, I790, I792, K551, K558, K559, Z958, Z959 |
| Cerebrovascular disease | G45, G46, I60, I61, I62, I63, I64, I65, I66, I67, I68, I69, H340 |
| Dementia | F00, F01, F02, F03, G30, F051, G311 |
| Chronic pulmonary disease excluding asthma* | J40, J41, J42, J43, J44, J46, J47, J60, J61, J62, J63, J64, J65, J66, J67, I278, I279, J684, J701, J703 |
| Asthma* | J45 |
| Connective tissue disease rheumatic disease | M05, M32, M33, M34, M06, M315, M351, M353, M360 |
| Peptic ulcer disease | K25, K26, K27, K28 |
| Mild liver disease | B18, K73, K74, K700, K701, K702, K703, K709, K717, K713, K714, K715, K760, K762, K763, K764, K768, K769, Z944 |
| Diabetes | E100, E101, E106, E108, E109, E110, E111, E116, E118, E119, E120, E121, E126, E128, E129, E130, E131, E136, E138, E139, E140, E141, E146, E148, E149, E102, E103, E104, E105, E107, E112, E113, E114, E115, E117, E122, E123, E124, E125, E127, E132, E133, E134, E135, E137, E142, E143, E144, E145, E147 |
| Paraplegia and hemiplegia | G81, G82, G041, G114, G801, G802, G830, G831, G832, G833, G834, G839 |
| Renal disease | N18, N19, N052, N053, N054, N055, N056, N057, N250, I120, I131, N032, N033, N034, N035, N036, N037, Z490, Z491, Z492, Z940, Z992 |
| Metastatic carcinoma | C77, C78, C79, C80 |
| Cancer | C00, C01, C02, C03, C04, C05, C06, C07, C08, C09, C10, C11, C12, C13, C14, C15, C16, C17, C18, C19, C20, C21, C22, C23, C24, C25, C26, C30, C31, C32, C33, C34, C37, C38, C39, C40, C41, C43, C45, C46, C47, C48, C49, C50, C51, C52, C53, C54, C55, C56, C57, C58, C60, C61, C62, C63, C64, C65, C66, C67, C68, C69, C70, C71, C72, C73, C74, C75, C76, C81, C82, C83, C84, C85, C88, C90, C91, C92, C93, C94, C95, C96, C97 |
| Moderate or severe liver disease | K704, K711, K721, K729, K765, K766, K767, I850, I859, I864, I982 |
| Aids (HIV) | B20, B21, B22, B24 |
| Hypertension* | I10, I11, I12, I13, I15 |

\* These disease groups have been changed from what is proposed in (Quan, et al., 2005). The two changes are splitting the “Chronic pulmonary disease” group into “Chronic pulmonary disease excluding asthma” and “Asthma” and adding the “Hypertension” group.

### Analysis of association of medications with COVID-19 presentation and mortality

We investigated the association of pharmacological therapy with COVID-19 positive presentation and inpatient mortality in COVID-19 patients. To assess the association of a given medication with these end points, we utilised the same stringent statistical approach that was described above for the comorbidities analysis, including matching on age, sex, and number of admissions, as well as the robustness analysis with different proportions of the full dataset (Figure 1). However, two further aspects were considered in this analysis; the first was conditioning on comorbidities; that is, the association of each medication with end points was assessed separately for patients with different comorbidities (Figure 2). This is to reduce the possibility of confounding observed association of medications with outcomes by their association with diseases. The definitions of comorbidity ICD-10 codes were based on Table 1.

**Figure 2.**
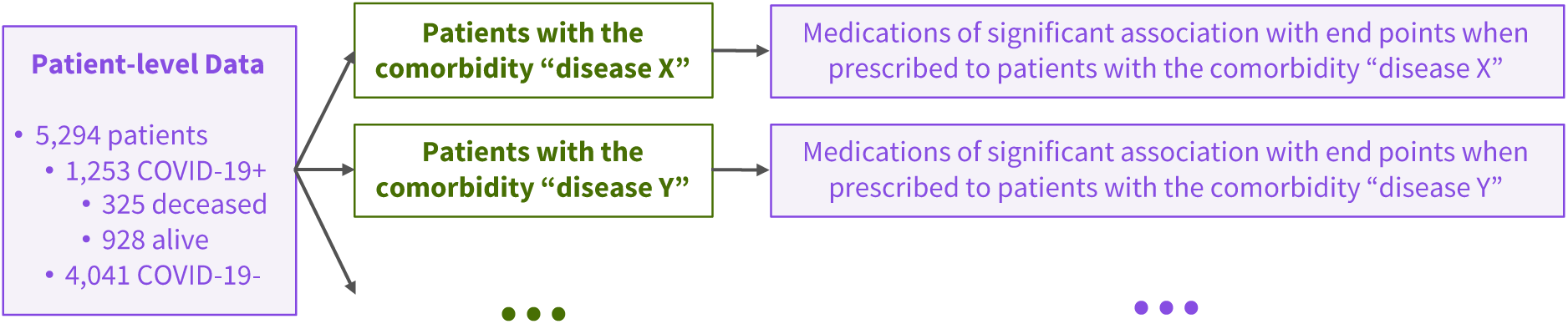
Analysis of medications was performed after conditioning on comorbidities.

The second aspect was the incorporation of the time of prescription. This is because the effects of medications can be dependent on the time of admission. Figure 3 illustrates the six lines of analysis that were conducted separately for each medication conditioned on comorbidities. The lines of analysis were designed to handle short-term and long-term medications differently. We define a short-term medication as a medication that was prescribed to the patient within the short period of time of indicated in Figure 3 but never before that (Figure 3: PP-ST1, PP-ST2, M-ST1, and M-ST2). As for long-term medications, they are defined as those which were prescribed at least twice in the patient history, one of which is more recent (Figure 3: PP-LT and M-LT). In all of these lines of analysis, medications with no more than ten patients in their active arm were skipped.

**Figure 3.**
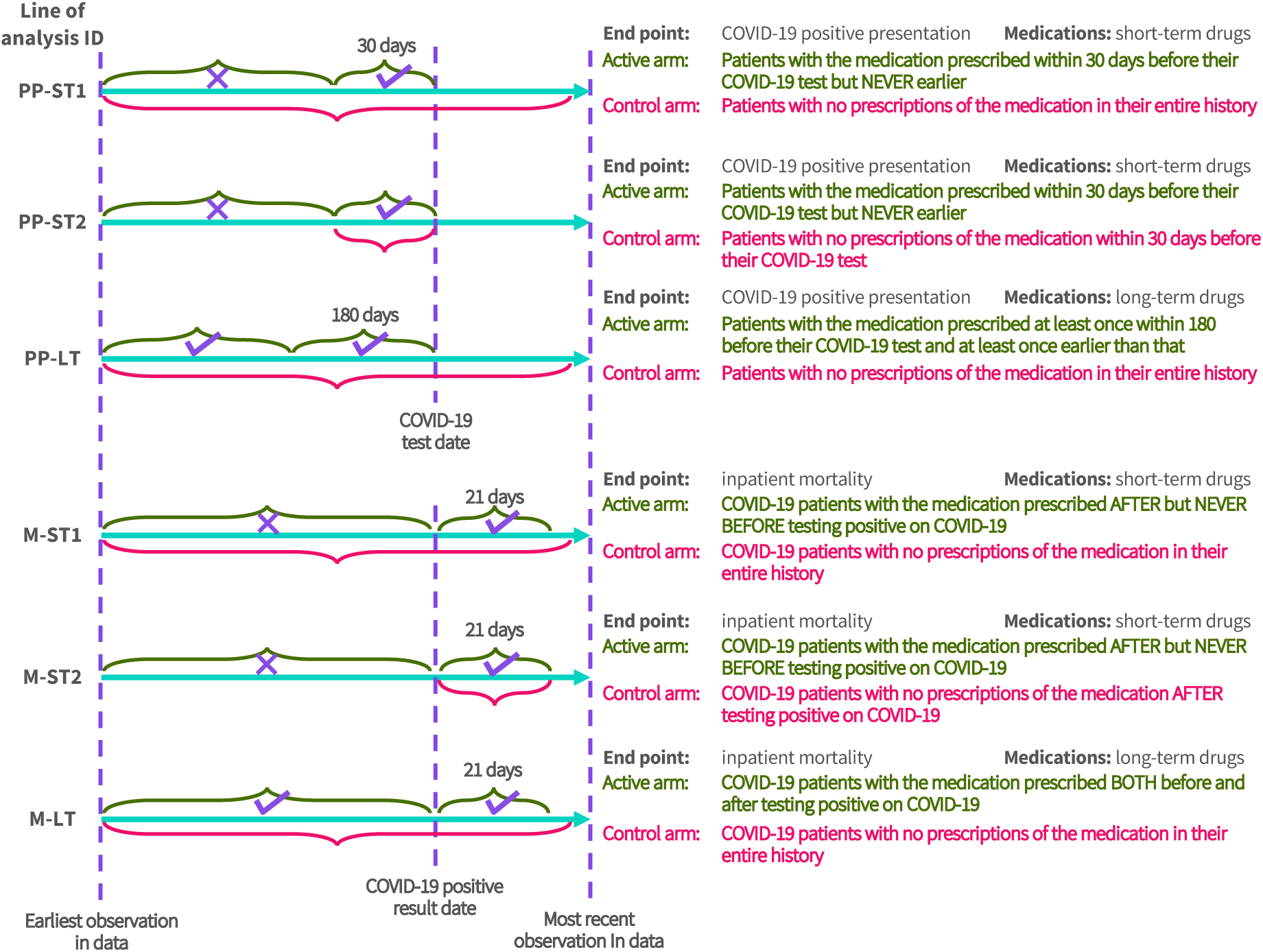
Lines of analysis of medications’ association with COVID-19 positive presentation or inpatient mortality in COVID-19 patients. The identifiers (IDs) for these lines of analysis are coded by a reference to the considered end point (PP: COVID-19 positive presentation; M: mortality) and a reference to the temporal consideration of the tested medications (ST: short-term; LT: long-term). If two lines of analysis share the same end point and temporal consideration, they are numbered with ordinal numbers.

Medications were encoded using British National Formulary (BNF) codes. BNF codes form a hierarchy where medications are classified into *BNF chapters*, which are sub-classified into *BNF sections*, which in turn are classified into *BNF paragraphs*, and so on up to eight levels of depth. In most of the cases, medications in our dataset are recorded with codes up to three or four levels of depth. When counting data records of a given BNF code, all medications of codes in deeper hierarchy levels are counted as well.

## Results

### Characteristics of the dataset

Figure 4 shows that the COVID-19 positive cohort has higher median age, more males, and patients with more hospital admissions than the COVID-19 negative cohort. A similar trend also appears in the cohort of deceased COVID-19 patients compared to the cohort of COVID-19 patients with no observed death in hospital (Figure 4).

**Figure 4.**
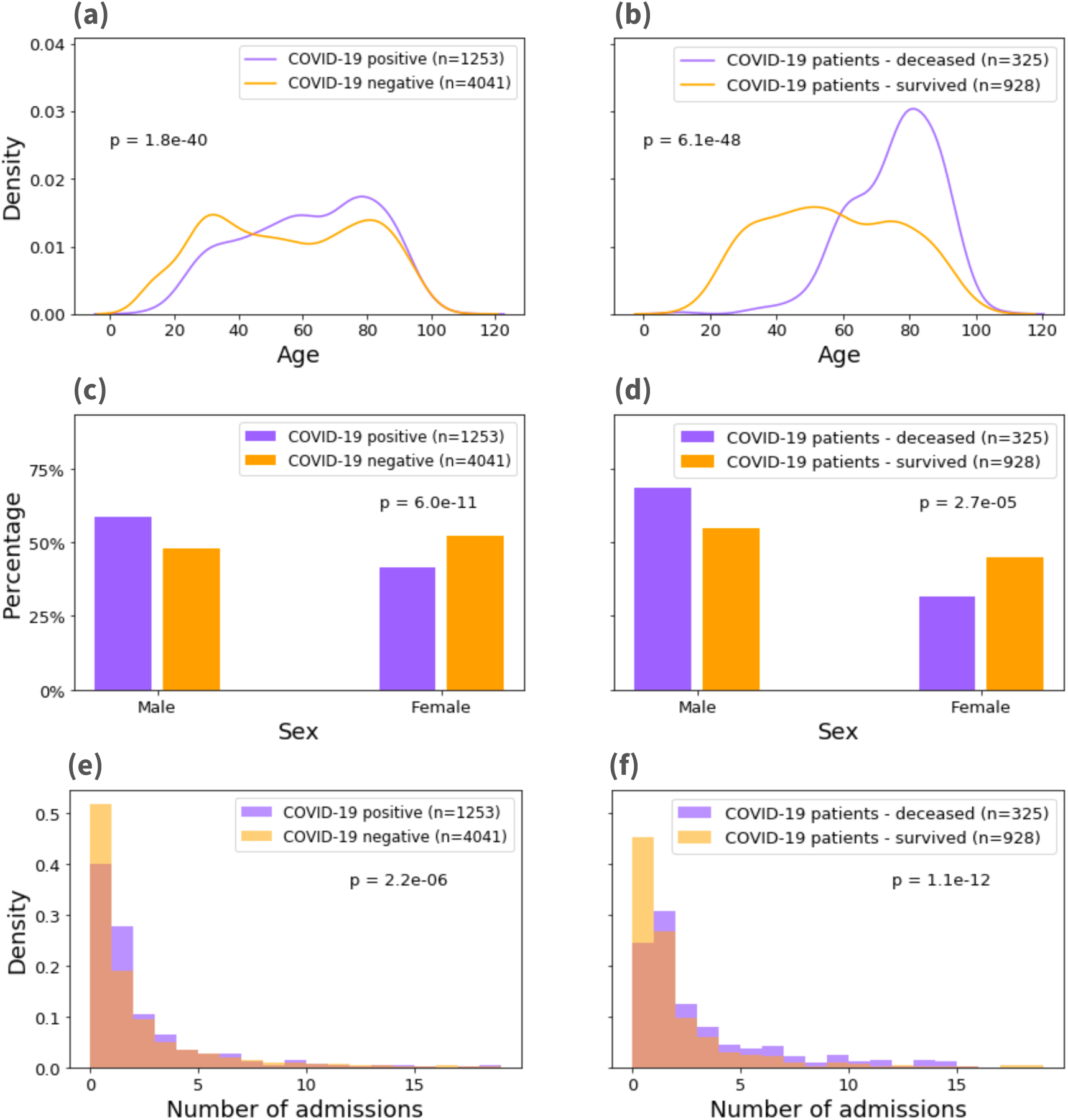
Distributions of age (a, b), sex (c, d), and number of admissions in patient’s history (e, f), each in COVID-19 positive versus negative patients (a, c, e), and in COVID-19 positive deceased versus survived patients (b, d, f). Calculation of p values was done using the unpaired two-sided Wilcoxon rank-sum test for age and number of admissions distributions (a, b, e, f), and using the two-sided Fisher’s exact test for sex distribution (c, d).

### Association of comorbidities with COVID-19 presentation and mortality

We observed that the comorbidities of hypertension and diabetes have statistically significant associations with the diagnosis of COVID-19 (Table 2). Moreover, hypertension, diabetes, congestive heart disease, and renal disease show significant association with higher inpatient mortality in COVID-19 patients (Table 3). These results were obtained after controlling for the effect of age, sex, and number of admissions in patients’ history.

**Table 2.**
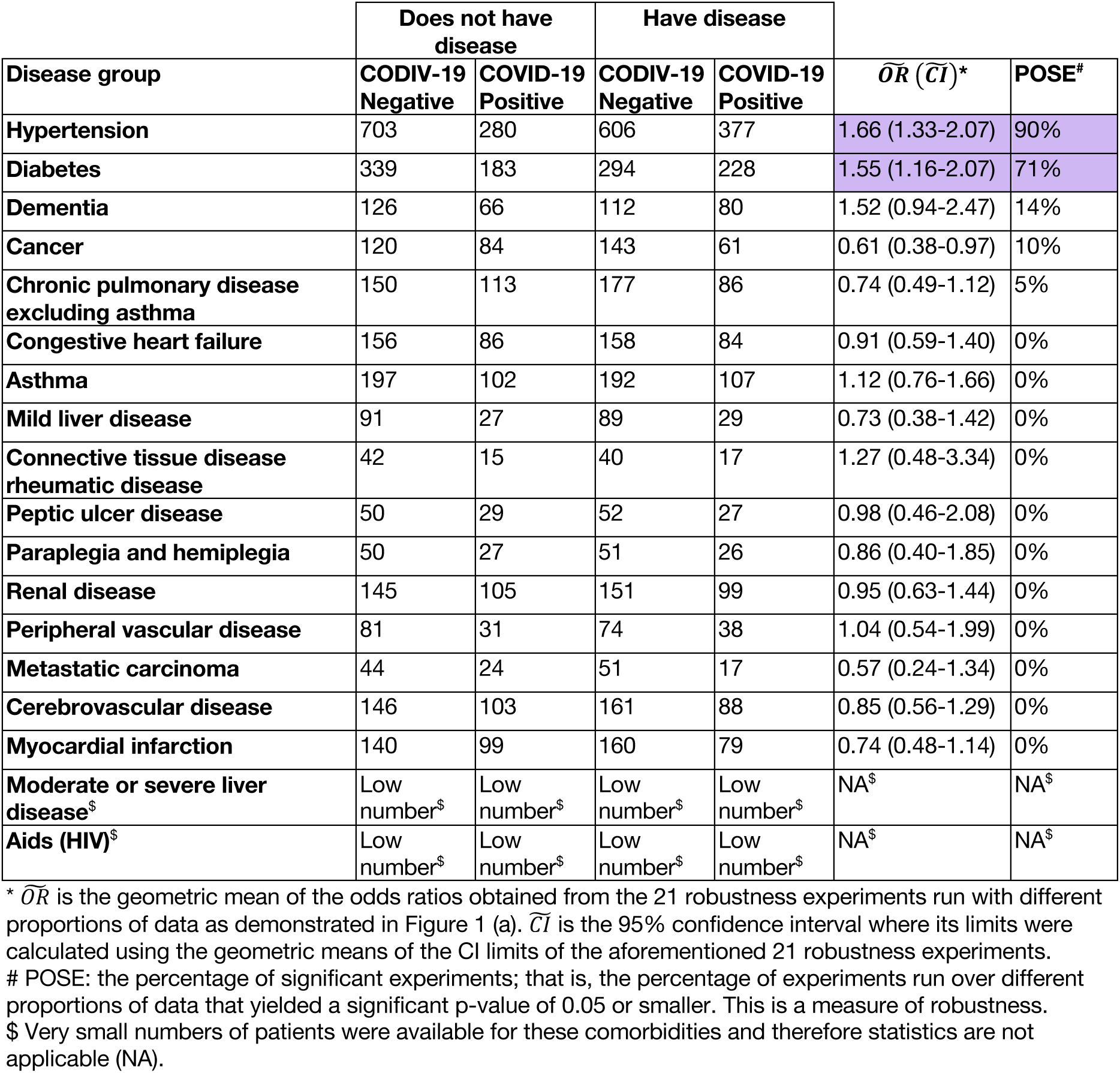
Comorbidity association with presentation with COVID-19 positive diagnosis

**Table 3.** Comorbidity association with mortality of COVID-19 positive patients

| Disease group | Does not have disease | | Have disease | | $\bar{OR} (\bar{CI})^*$ | POSE<sup>#</sup> |
| --- | --- | --- | --- | --- | --- | --- |
|  | Survived | Deceased | Survived | Deceased |  |  |
| Hypertension | 294 | 83 | 214 | 163 | 2.70 (1.87-3.92) | 100% |
| Diabetes | 176 | 52 | 127 | 101 | 2.20 (1.38-3.49) | 95% |
| Congestive heart failure | 57 | 27 | 35 | 49 | 3.05 (1.45-6.41) | 81% |
| Renal disease | 61 | 38 | 40 | 59 | 2.63 (1.35-5.13) | 76% |
| Cerebrovascular disease | 59 | 29 | 44 | 44 | 2.09 (1.02-4.29) | 38% |
| Myocardial infarction | 47 | 32 | 35 | 44 | 2.13 (1.01-4.49) | 24% |
| Peripheral vascular disease | 27 | 11 | 17 | 21 | 3.29 (1.04-10.4) | 19% |
| Dementia | 47 | 33 | 35 | 45 | 1.89 (0.90-3.96) | 14% |
| Metastatic carcinoma | 10 | 7 | 5 | 12 | 4.56 (0.79-26.3) | 10% |
| Cancer | 36 | 25 | 31 | 30 | 1.61 (0.69-3.76) | 5% |
| Peptic ulcer disease | 16 | 11 | 14 | 13 | 1.61 (0.44-5.87) | 5% |
| **Chronic pulmonary disease excluding asthma** | 46 | 40 | 43 | 43 | 1.65 (0.81-3.36) | 0% |
| **Asthma** | 73 | 34 | 69 | 38 | 1.20 (0.62-2.32) | 0% |
| **Mild liver disease** | 22 | 7 | 20 | 9 | 1.04 (0.27-3.98) | 0% |
| **Paraplegia and hemiplegia** | 16 | 10 | 16 | 10 | 1.50 (0.38-5.88) | 0% |
| **Connective tissue disease rheumatic disease** | 10 | 7 | 12 | 5 | 0.57 (0.10-3.24) | 0% |
| **Moderate or severe liver disease<sup>§</sup>** | Low number<sup>§</sup> | Low number<sup>§</sup> | Low number<sup>§</sup> | Low number<sup>§</sup> | NA<sup>§</sup> | NA<sup>§</sup> |
| **Aids (HIV)<sup>§</sup>** | Low number<sup>§</sup> | Low number<sup>§</sup> | Low number<sup>§</sup> | Low number<sup>§</sup> | NA<sup>§</sup> | NA<sup>§</sup> |

\* $\overline{OR}$ is the geometric mean of the odds ratios obtained from the 21 robustness experiments run with different proportions of data as demonstrated in Figure 1 (a). $\overline{CI}$ is the 95% confidence interval where its limits were calculated using the geometric means of the CI limits of the aforementioned 21 robustness experiments.

### Association of medications with COVID-19 presentation and mortality

Table 4 shows the numbers of medications qualified for testing in each line of analysis given that they were prescribed to more than 10 patients within the relevant time window (Figure 3). This table also shows the number of medications, out of all of those qualified for testing, that showed significant and robust association with reduced presentation with COVID-19 (lines PP-ST1, PP-ST2, and PP-LT) or with reduced inpatient mortality in COVID-19 patients (lines M-ST1, M-ST2, and M-LT).

**Table 4.** Summary of numbers of medications qualified for testing in each one of the six lines of analysis and numbers of medications with significant association with reduced COVID-19 positive presentation or with reduced mortality in COVID-19 patients.

| Line of analysis ID | Medications qualified for testing (those with more than 10 patients in their active arm) |  |  | Medications associated with reduced COVID-19 positive presentation (PP- lines) or with reduced mortality (M- lines) |  |  |
| --- | --- | --- | --- | --- | --- | --- |
|  | BNF Sections | BNF paragraphs | BNF sub-paragraphs | BNF Sections | BNF paragraphs | BNF sub-paragraphs |
| PP-ST1 | 21 | 18 | 5 | 0 | 0 | 0 |
| PP-ST2 | 22 | 19 | 5 | 0 | 0 | 0 |
| PP-LT | 112 | 53 | 10 | 0 | 0 | 0 |
| M-ST1 | 53 | 37 | 10 | 10 | 7 | 2 |
| M-ST2 | 50 | 36 | 10 | 11 | 6 | 1 |
| M-LT | 60 | 27 | 5 | 1 | 0 | 0 |

None of the tested medications was significantly and robustly associated with reduced COVID-19 presentation in our results, whether while considering short-term medications (PP-ST1 and PP-ST2) or long-term medications (PP-LT). Results with different thresholds of robustness are provided in Supplementary File S1.

Table 5 lists medications that passed the test of robust statistical significance. Where a BNF section and one of its BNF paragraphs both appear significant, the one that is less specific is omitted from this table for a more concise display. Nonetheless, all tested medications are listed in Supplementary File S2.

**Table 5.** Statistically significant and robust associations between medications and mortality for COVID-19+ patients. The full list of tested medications is included in Supplementary File S1.

|  |  | Not on Medication |  | On Medication |  |  |  |
| --- | --- | --- | --- | --- | --- | --- | --- |
| Disease group | Drug BNF code / drug name | Survived | Deceased | Survived | Deceased | $\widehat{OR}(\widehat{CI})^*$ | POSE<sup>#</sup> |
| Intersection of M-ST1 and M-ST2<sup>s</sup> |  |  |  |  |  |  |  |
| Diabetes | 02.08.02<br>Oral Anticoagulants | 72 | 96 | 21 | 3 | 0.11 (0.03-0.50) | 100% |
|  | 02.12<br>Lipid-Regulating Drugs | 70 | 90 | 22 | 4 | 0.15 (0.04-0.58) | 86% |
|  | 05.01.01.03<br>Broad-Spectrum Penicillins | 73 | 94 | 24 | 5 | 0.20 (0.06-0.64) | 90% |
|  | 06.01.02.02<br>Biguanides | 74 | 94 | 19 | 4 | 0.19 (0.05-0.70) | 81% |
| Hypertension | 01.03<br>Antisecretory Drugs+Mucosal Protectants | 120 | 148 | 31 | 9 | 0.22 (0.09-0.56) | 100% |
|  | 01.06<br>Laxatives | 129 | 153 | 21 | 3 | 0.10 (0.02-0.45) | 95% |
|  | 02.05.05.01<br>Angiotensin-Converting Enzyme Inhibitors | 140 | 160 | 17 | 1 | 0.06 (0.01-0.53) | 81% |
|  | 02.06.02<br>Calcium-Channel Blockers | 122 | 154 | 33 | 5 | 0.12 (0.04-0.36) | 100% |
|  | 02.08.01<br>Parenteral Anticoagulants | 144 | 160 | 19 | 2 | 0.09 (0.02-0.50) | 90% |
|  | 02.08.02<br>Oral Anticoagulants | 122 | 153 | 27 | 3 | 0.08 (0.02-0.35) | 100% |
|  | 09.02<br>Antiplatelet Drugs | 139 | 158 | 16 | 2 | 0.10 (0.02-0.59) | 95% |
|  | 02.12<br>Lipid-Regulating Drugs | 118 | 148 | 33 | 6 | 0.15 (0.05-0.46) | 95% |
|  | 04.07.01<br>Non-Opioid Analgesics And Compound Prep | 136 | 152 | 19 | 5 | 0.22 (0.07-0.69) | 76% |
|  | 05.01.01.03<br>Broad-Spectrum Penicillins | 131 | 156 | 29 | 5 | 0.14 (0.05-0.45) | 100% |
| M-ST1 other than those that also appeared in M-ST2 |  |  |  |  |  |  |  |
| None |  |  |  |  |  |  |  |
| M-ST2 other than those that also appeared in M-ST1 |  |  |  |  |  |  |  |
| Diabetes | 02.05<br>Hypertension and Heart Failure | 79 | 92 | 14 | 3 | 0.15 (0.03-0.66) | 71% |
|  | 02.06<br>Nit,Calc Block & Other Antianginal Drugs | 79 | 96 | 14 | 2 | 0.11 (0.02-0.60) | 86% |
| Hypertension | 02.02<br>Diuretics | 134 | 151 | 21 | 8 | 0.33 (0.12-0.91) | 81% |
|  | 02.04<br>Beta-Adrenoceptor Blocking Drugs | 134 | 152 | 19 | 7 | 0.29 (0.10-0.86) | 86% |
| M-LT |  |  |  |  |  |  |  |
| Diabetes | 06.01<br>Drugs Used In Diabetes | 30 | 65 | 26 | 12 | 0.26 (0.10-0.67) | 71% |

\* $\bar{OR}$ is the geometric mean of the odds ratios obtained from the 21 robustness experiments run with different proportions of data as demonstrated in Figure 1 (a). $\bar{CI}$ is the 95% confidence interval where its limits were calculated using the geometric means of the CI limits of the aforementioned 21 robustness experiments.

Considering short-term medications (M-ST1 & M-ST2), we observed an association with lower COVID-19 mortality for both diabetic and hypertensive patients for whom anticoagulants, lipid-regulating drugs, and penicillins were administered after their positive COVID-19 test (Table 5).

Biguanides prescribed to diabetic patients after testing positive on COVID-19 were associated with reduced mortality (Table 5). Interestingly, the more general BNF section of drugs used in diabetes, which includes insulins as well as antidiabetic drugs, showed association with reduced COVID-19 mortality for diabetic patients for whom there is an evidence of prescription before and after a positive COVID-19 test (M-LT) (Table 5).

Furthermore, angiotensin-converting enzyme inhibitors (ARBs) and antiplatelet drugs showed association with reduced COVID-19 mortality when administered to hypertensive patients after testing positive on COVID-19 (M-ST1 & M-ST2) (Table 5).

## Discussion

We describe the retrospective analysis of 5294 patients presenting to two hospitals in London; 1253 of whom were COVID-19 positive. We observe that having a diagnosis of hypertension or diabetes was associated with a higher presentation with COVID-19, and that hypertension, diabetes, congestive heart failure, and renal disease were associated with a higher chance of in-hospital death for COVID-19 positive patients. Furthermore, we observe associations between a number of medications prescribed after a positive COVID-19 test and reduced mortality; examples include anticoagulants, lipid-regulating drugs, and penicillins for diabetic and hypertensive patients, biguanides for diabetic patients, and ARBs and antiplatelets drugs for hypertensive patients. We also observed an association with reduced COVID-19 mortality for diabetic patients who have been on diabetic drugs even before testing positive on COVID-19.

Co-morbidities associated with COVID-19 have been reported previously in the literature for a number of diseases such as hypertension (Williams & Zhang, 2020; Vizcaychipi, et al., 2020), renal diseases, and diabetes (Wu & McGoogan, 2020; Chen, et al., 2020; Yang, et al., 2020; Wang, et al., 2020; Perico, et al., 2020; Vizcaychipi, et al., 2020). COVID-19 patients with such co-morbidities were more likely to present to hospital which is in line with many NHS triage systems who recognise this group as vulnerable and therefore are more likely to recommend hospital attendance.

Some of our observations of associations of medications are in-line with published literature, such as the protective effect of anticoagulants patients on COVID-19 outcomes (Atallah, et al., 2020; Tang, et al., 2020). On the other hand, some of our observations on medications were at odds with the published literature or published literature gave conflicting results, such as the effect of ARBs (Perico, et al., 2020; Cure & Cure, 2020a; Cure & Cure, 2020b).

For all lines of analyses of comorbidities and medications, matching active and control arms on age, sex, and number of admissions was essential to account for substantial confounding factors. The essence of this is indicated by observing that the distributions of these covariates differed significantly between patients who had positive and negative COVID-19 test results as well as between COVID-19 patients who died and who did not (Figure 4). Matching the number of hospital admissions aims at comparing patients with more comparable medical history and overall health condition.

### Limitations

Observed associations should be interpreted with care as they might be attributable to confounding factors. For example, we did not use measures of functional status, e.g. clinical fragility scale, which may have been over-represented in one arm of our analysis. Furthermore, we have not considered interactions between multiple co-morbidities and their association with outcomes. Severity of COVID-19 infection at presentation was not incorporated and no distinction is being made in this analysis between those who have been admitted for an acute episode of some comorbidity in their most recent admission and those who have not despite having that condition chronically. Also, length of symptoms at point of presentation to hospital were not assessed neither was the frailty scale.

Observations in this study are based on a population of patients presenting to hospitals and for whom COVID-19 status has been assessed to be positive or negative. This may cause a bias as it is not comparing individuals belonging to the general community population. Also, this population of patients comes from two London metropolitan hospitals that may not have a similar distribution of comorbidities and features as hospitals in other metropolitan cities or hospitals away from large cities. Generalisability is therefore not assured without further confirmatory studies. Additionally, our data does not include medications prescribed in primary care, resulting in a potentially inaccurate representation of the medication history of patients, especially prior to having a positive COVID-19 test result.

Finally, low data coverage and bias may cause absence of statistically significant associations with COVID-19 outcomes for some comorbidities or medications. Therefore, no conclusions may be drawn for such cases without further investigation.

### Conclusions

This study provides an important piece of real-world evidence on associations between co-morbidities and medication prescription, respectively with COVID-19 positivity presenting to hospital and inpatient mortality. Identifying these associations can help in the crucial task of defining the vulnerable groups that may benefit from more stringent social distancing especially as lockdown due to COVID-19, is relaxed. Nonetheless, observations in this study have to be interpreted with caution due to potential bias and confounders, and confirmatory studies will be required to draw reliable conclusions.

## Data Availability

_

## Acknowledgements

This work uses data provided by patients and collected by the NHS as part of their care and support. We believe using patient data is vital to improve health and care for everyone and would, thus, like to thank all those involved for their contribution.

Special thanks are due to the Chelsea and Westminster (CW) COVID-19 AICU Consortium, comprising all critical care personnel who were part of the delivery of care during the COVID444 19 pandemic as follows: CW Anaesthetics Consultants, Critical Care Consultants, Trainees & Fellows from ICU, Anaesthesia, and seconded to ICU from other specialities, Surgeons, the supporting Respiratory and ED Physicians, Operating Department Practitioners and CW Critical Care Nurses. This united approach to an unprecedented clinical condition was critical not only to the management of the patients but also to our ability to document and collate the key data in a timely manner to support this analysis.

Also, a special thank you to Trystan Hawkin, Chris Chaney from CWplus, the Planned Care Clinical Division managers, porters, domestic personnel and the CW local community who without hesitation have supported the National Healthcare System throughout the COVID-19 pandemic.

## Compliance with ethical guidelines

All methods were performed in accordance with the relevant guidelines and regulations.

The study was approved by the IG management team of Sensyne Health plc and Chelsea & Westminster NHS Foundation Trust under the Strategic Research Agreement (SRA) and relative Data Sharing Agreements (DSAs) signed by the NHS Trust and Sensyne Health plc on 25th July 2018.

All analyses were conducted on data with no personal identifying information. Therefore, informed consent was waived by the ethics committee of the Chelsea & Westminster NHS Foundation Trust, which provided ethical approval for the study.

## Notes

### Competing Interest Statement

BAJ, AA, JB, LM, and RTK are employees of Sensyne Health plc (part-time in the case of LM) or were employed by Sensyne Health plc over the time period in which they contributed to this study. LM is further funded by The National Institute for Health Research Grant (IS-BRC-1215-20008).

### Funding Statement

The study was supported by Sensyne Health plc. LM is further funded by The National Institute for Health Research Grant (IS-BRC-1215-20008). The funders had no role in study design, data collection and analysis, decision to publish, or preparation of the manuscript

### Author Declarations

The study was approved by the IG management team of Sensyne Health plc and Chelsea & Westminster NHS Foundation Trust under the Strategic Research Agreement (SRA) and relative Data Sharing Agreements (DSAs) signed by the NHS Trust and Sensyne Health plc on 25th July 2018. All analyses were conducted on data with no personal identifying information. Therefore, informed consent was waived by the ethics committee of the Chelsea & Westminster NHS Foundation Trust, which provided ethical approval for the study.

